# Deep learning identified genetic variants associated with COVID-19 related mortality

**DOI:** 10.1101/2022.05.05.22274731

**Authors:** Zihuan Li, Wei Dai, Shiying Wang, Yisha Yao, Heping Zhang

**Affiliations:** School of Public Health, Yale University, New Haven, CT, USA

**Author notes:** Corresponding author: Heping Zhang. 60 College Street, New Haven, CT, 06520-8034, USA.

**Keywords:** Deep learning, COVID-19, SARS-CoV-2, UK Biobank, TAS2R1

## Abstract

Analysis of host genetic components provides insights into the susceptibility and response to viral infection such as severe acute respiratory syndrome coronavirus 2 (SARS-CoV-2), which causes coronavirus disease 2019 (COVID-19). To reveal genetic determinants of susceptibility to COVID-19 related mortality, we train a deep learning model to identify groups of genetic variants and their interactions that contribute to the COVID-19 related mortality risk using the UK Biobank data. We refer to such groups of variants as super variants. We identify 15 super variants with various levels of significance as susceptibility loci for COVID-19 mortality. Specifically, we identify a super variant (OR=1.594, *p*=5.47×10^−9^) on Chromosome 7 that consists of the minor allele of rs76398985, rs6943608, rs2052130, 7:150989011_CT_C, rs118033050 and rs12540488. We also discover a super variant (OR=1.353, *p*=2.87×10^−8^) on Chromosome 5 that contains rs12517344, rs72733036, rs190052994, rs34723029, rs72734818, 5:9305797_GTA_G and rs180899355.

## Introduction

Coronavirus disease 2019 (COVID-19) is a pandemic viral disease caused by the severe acute respiratory syndrome coronavirus 2 (SARS-CoV-2) and has resulted in significant morbidity and mortality worldwide.

The ongoing research efforts to provide new insights into risk of COVID-19 related morbidity and mortality have focused largely on investigating the epidemiological characteristics of COVID-19 [1, 2], and genomic characterization of COVID-19 [3]. According to a cross-sectional survey conducted in the United Kingdom, among the hospitalized COVID-19 patients, those with diabetes, cardiovascular diseases, kidney diseases, obesity, or chronic respiratory diseases are tied to high risk of death [4, 5]. In addition, several recent papers have revealed that males, older people, Black, Asian and Minority Ethnic groups are exposed to an elevated risk of COVID-19 caused mortality [6-9]. Recent evidence also suggests that host genetic determinants modulate the risk of infection and disease severity [10-13]. Thus, investigating the host genetic basis of heterogeneous susceptibility and severity of COVID-19 and identifying genetic risk factors will deepen our understanding and facilitate the drug developments of COVID-19.

Several genome-wide association studies (GWAS) have been performed to investigate the genetic contribution to COVID-19 outcomes, including the associations between host genetic factors, specifically single-nucleotide polymorphisms (SNPs) and infection or respiratory failure [4, 10, 13, 14], but those studies only focus on the effects of individual SNPs on phenotypes. While most of the literature mentioned above investigate the effects of the variants on COVID-19 susceptibility and severity, few examined association between SNPs and COVID-19 mortality. It is important to examine interactions between SNPs or genes on COVID-19 mortality. Several works utilizing tree-based method have been proposed to overcome drawbacks of traditional GWAS [15-18]. For example, Hu et al. combined a local ranking and aggregation approach with random forest to identify genetic variants associated with risk of COVID-19 related mortality [17]. Though they demonstrated some promising results, including the discovery of some new interacting SNPs, they used an earlier release of the UK Biobank data which contained a much smaller sample size on the COVID-19 cases and deaths.

To account for the potential interactions between SNPs or genes, we adopt the idea of super variants in Song et al [16]. A super variant is a combination of minor alleles in multiple loci throughout a genome in analog to a gene [16]. In contrast to a gene, which is a physically connected region on a chromosome, the loci contributing to a super variant might be located anywhere in the genome. Recent studies have demonstrated the usefulness of super variants in detecting genetic associations with complex diseases [15, 17, 18]. To incorporate potentially complex interactions more effectively among SNPs or genes, we propose a novel approach, namely deep learning-based ranking and aggregation method for identifying genetic variants (DRAG).

To date, deep learning has been successfully applied to numerous tasks in the biomedical domain, such as genomics [19], promoters [20]. Although deep learning methods have been applied in GWAS [21], those methods do not emphasize SNP interaction effects. By contrast, our proposed method is capable of detecting the main and interaction effects of SNPs that are entangled in high-dimensional and nonlinear representations. In addition, deep learning-based feature selection approaches such as Concrete Auto-encoder (CAE) have been studied by [22, 23]. Abid et al. demonstrated the advantages of CAE: 1) efficacy on a large-scale gene expression dataset; 2) easily scales to high-dimensional datasets; and 3) outperforms a variety of other sophisticated feature selection approaches, including principal feature analysis, multi-cluster feature selection and auto-encoder inspired unsupervised feature selection [23].

The DRAG proceeds in three main steps: SNP-set partition, selecting an optimal subset of SNPs, and determination of super variants. In the first step, SNPs are first divided into consecutive non-overlapping regions with each region consisting multiple SNPs. In the next step, an optimal subset of SNPs within each SNP-set is selected using CAE combined with Bayesian information criterion (BIC) obtained by training logistic regression. Finally, a super variant is formed by the selected subset of SNPs, and the association between the super variant and phenotype is tested by performing logistic regression.

We first apply the DRAG to identify super variants that contribute to the COVID-19 related mortality risk using the UK Biobank white British data to minimize the race and ethnicity impact on our results. We include 18,731 with 8.2 million SNPs as the discovery set. The identified super variants are then tested in an independent validation dataset (n=9,366). Furthermore, we demonstrate the validity and efficacy of our analytic approach through simulation studies.

## Results

### DRAG identifies super variants associated with COVID-19 related mortality

We include a total of 28,097 individuals infected with COVID-19 (26,441 survived and 1,656 died) and of white British ancestry from the UK Biobank in our analyses. We consider 8,238,098 SNPs which are divided into 2734 SNP-sets. Each SNP-set is 1Mbp as done in Hu et al. [16, 17] for convenience.

The discovery set contains about two thirds of the participants which lead to the identification of 15 super variants with p-values at or below 1.83×10^−5^ (i.e., 0.05/2734) (Fig. 1).

**Figure 1:**
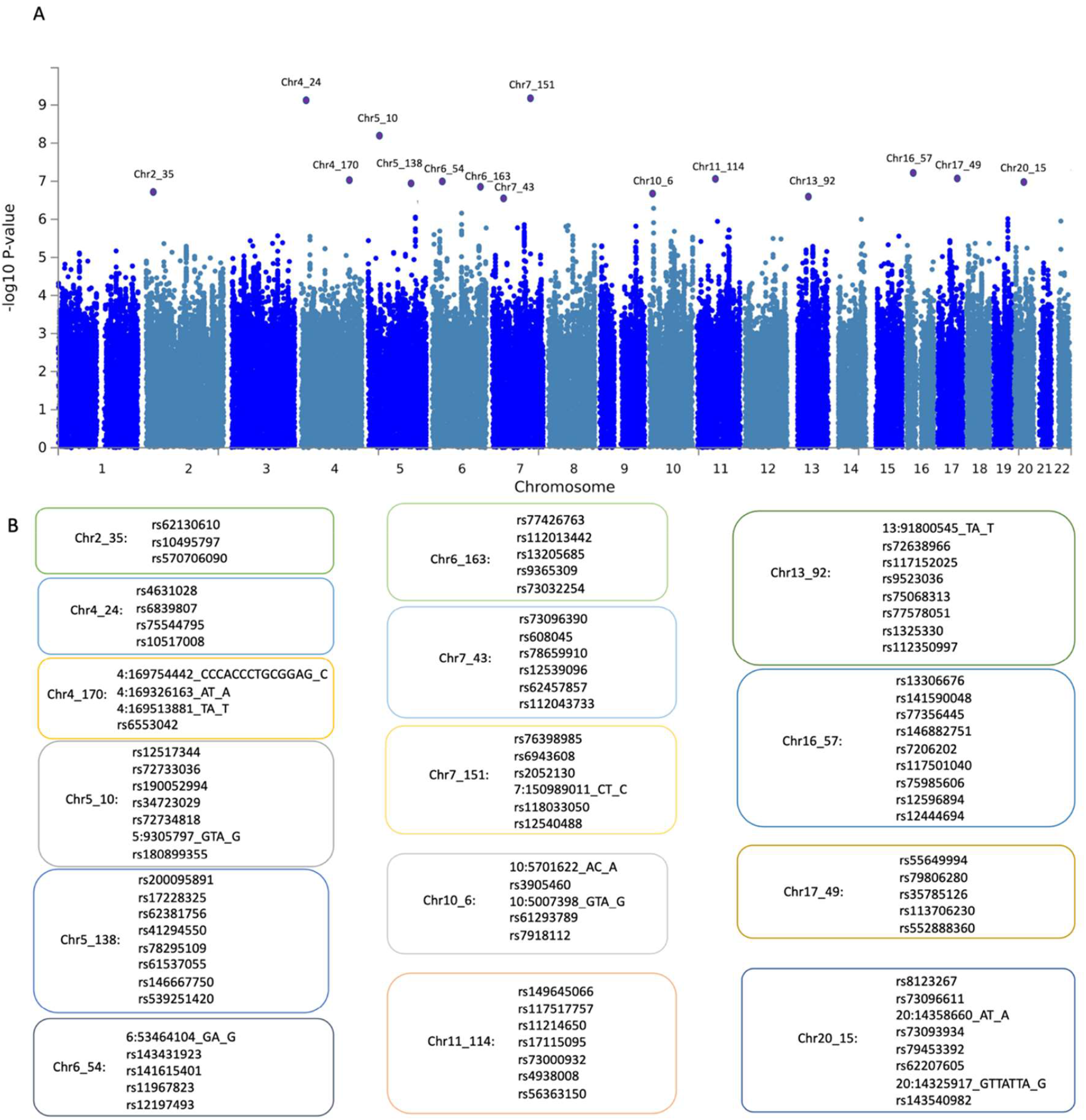
**(A)** Manhattan plot. **(B)** The set of SNPs in each super variant. The chri_j represents the super variant in the *j-*th set of the *i-*th chromosome.

As the next step, we validate the detected super variants in the remaining one third of the individuals. All of the 15 detected super variants have p-values below 0.05 and one below 0.003 (0.05/15) (Table 1).

**Table 1:** Marginal effects of 15 verified super variants on the discovery, verification, and complete dataset using DRAG. The chri_j represents the super variant in the *j-*th set of the *i-*th chromosome; OR on discovery: odd ratio of super variant on discovery set; OR on verification: odd ratio of super variant on verification set; OR on complete: odd ratio of variant on complete set.

| Supervariants | OR on discovery | p-value on discovery | OR on verification | p-value on verification | OR on complete | p-value on complete |
| --- | --- | --- | --- | --- | --- | --- |
| chr2_35 | 1.401 | $7.51 \times 10^{-6}$ | 1.289 | $1.87 \times 10^{-2}$ | 1.368 | $4.51 \times 10^{-7}$ |
| chr4_24 | 1.464 | $7.33 \times 10^{-9}$ | 1.214 | $3.58 \times 10^{-2}$ | 1.376 | $2.40 \times 10^{-9}$ |
| chr4_170 | 1.381 | $6.11 \times 10^{-6}$ | 1.303 | $9.01 \times 10^{-3}$ | 1.354 | $2.18 \times 10^{-7}$ |
| chr5_10 | 1.343 | $1.01 \times 10^{-5}$ | 1.383 | $6.07 \times 10^{-4}$ | 1.353 | $2.87 \times 10^{-8}$ |
| chr5_138 | 1.361 | $2.59 \times 10^{-6}$ | 1.207 | $4.23 \times 10^{-2}$ | 1.306 | $5.71 \times 10^{-7}$ |
| chr6_54 | 1.395 | $5.41 \times 10^{-7}$ | 1.212 | $4.24 \times 10^{-2}$ | 1.329 | $1.67 \times 10^{-7}$ |
| chr6_163 | 1.439 | $1.04 \times 10^{-5}$ | 1.342 | $1.31 \times 10^{-2}$ | 1.406 | $5.03 \times 10^{-7}$ |
| chr7_43 | 1.398 | $1.61 \times 10^{-5}$ | 1.329 | $1.12 \times 10^{-2}$ | 1.370 | $7.61 \times 10^{-7}$ |
| chr7_151 | 1.688 | $6.74 \times 10^{-8}$ | 1.419 | $1.37 \times 10^{-2}$ | 1.594 | $5.47 \times 10^{-9}$ |
| chr10_6 | 1.359 | $5.72 \times 10^{-6}$ | 1.217 | $4.27 \times 10^{-2}$ | 1.309 | $1.12 \times 10^{-6}$ |
| chr11_114 | 1.364 | $6.07 \times 10^{-6}$ | 1.242 | $2.68 \times 10^{-2}$ | 1.321 | $7.03 \times 10^{-7}$ |
| chr13_92 | 1.374 | $1.05 \times 10^{-5}$ | 1.226 | $4.94 \times 10^{-2}$ | 1.320 | $2.68 \times 10^{-6}$ |
| chr16_57 | 1.425 | $4.18 \times 10^{-6}$ | 1.341 | $6.98 \times 10^{-3}$ | 1.394 | $1.19 \times 10^{-7}$ |
| chr17_49 | 1.379 | $1.95 \times 10^{-6}$ | 1.209 | $4.31 \times 10^{-2}$ | 1.314 | $5.80 \times 10^{-7}$ |
| chr20_15 | 1.333 | $1.59 \times 10^{-5}$ | 1.267 | $1.13 \times 10^{-2}$ | 1.310 | $6.40 \times 10^{-7}$ |

Table 1 shows the effects of super variants estimated from logistic regression with sex, age and top 10 principal components (PCs) of the genomic markers [24-26] in the discovery set, as well as the results in the verification and complete sets. The most significant signal for complete set is given by chr4_24 (*p* = 2.40 × 10^−9^), and the largest odds ratio appears at chr7_151 (OR = 1.594). The specific formation of the 15 verified super variants and their nearby genes (genes are annotated with ANNOVAR) are given in Table S1 (Supplementary Table 1) where the odds ratio and corresponding *p* values are calculated on the complete dataset.

### Annotation of the identified super variants in their reported roles related to COVID-19

Our analyses identify 4 genetic variants (in/near *TAS2R1, ZBTB16, LINC01320* and *NCAM1*) [11, 27-31] that have been reported with COVID-19 outcomes.

First, it is worth noting that super variant chr11_114 (OR=1.321, *p*=7.03×10^−7^) is composed of 7 SNPs, including the variant rs17115095 in the intronic region of gene *NCAM1* which encodes a cell adhesion protein belonging to the immunoglobulin superfamily. The encoded protein is involved in cell-to-cell and cell-to-matrix interactions during development and differentiation [32]. As a binding partner of spike protein, previous investigations have revealed that there might exist molecular mimicry between *NCAM1* and the COVID-19 envelope protein [29, 30]. Furthermore, in the same super variant, we observe that the SNP rs73000932 is considered as an intronic variant for gene *ZBTB16* (also referred to as promyelocytic leukemia zinc finger). This gene plays a critical role in the function and development of immune system and may enhance T cell responses [33]. Recent studies have reported the upregulation of *ZBTB16* in the tears of COVID-19 patients [31]. These findings suggest that variations within *ZBTB16* is likely to have an effect on the risk of COVID-19 related mortality.

Second, the 7 SNPs in super variant ch5_10 (OR=1.353, *p*=2.87×10^−8^), including the SNP index rs180899355, and its nearby gene *TAS2R1* are mapped to chromosome 5p15. This gene encodes a member of the G protein-coupled receptor superfamily that is expressed by taste receptor cells of the tongue and palate epithelia. This intronless taste receptor gene encodes a 7-transmembrane receptor protein, functioning as a bitter taste receptor [34]. Studies have shown that individuals who reported experiencing weak or no bitter tastes were considerably more likely to test positive for COVID-19, to be hospitalized, and to be symptomatic for a longer duration [35]. In addition, recent studies have been conducted that reduced *TAS2R* expression may influence the response to Chloroquine. COVID-19 infected obese patients could respond differently to pharmacological therapy with Chloroquine, with side effects occurring more frequently as result of overdosage [27]. Moreover, the rs180899355 is in the same region as a variant rs148943015 in *TAS2R* gene, specifically rs148943015, which is reported to be associated with COVID-19 susceptibility and severity [11].

In addition, super variant chr2_35 (OR=1.368, *p*=4.51×10^−7^) includes SNP rs570706090 whose nearby gene is *LINC01320*. Of note, *LINC01320* expression is reported to be higher in COVID-19 tissue specimens of connecting tubule (CNT) and intercalated cell type A (IC-A) [28]. In addition, a GWAS study has shown that increased expression of *LINC01320* is associated with testing positive for SARS-CoV-2 [11].

### DRAG identifies novel genes with variants associated with COVID-19 mortality

We also identify 8 novel genes (*DDX60L, HSPA9, LASTR, GLI3, AGAP3, MACROD2, NUP93, ELOVL5*) may affect COVID-19 related mortality.

Super variant chr4_170 (OR=1.354, *p*=2.18×10^−7^) is composed of 4 SNPs. Variant 4:169326163_AT_A is located in the intronic region of gene *DDX60L* which encodes a protein that is a member of the DEAD-box (DDX) helicase family. Although the function of this gene has not been well characterized, it has been shown that *DDX60L* is involved in anti-viral immunity and acts as a cytosolic sensor of viral nucleic acids [36-38]. The proteins responsible for DDX-mediated hijacking mechanisms are highly conserved among coronaviruses [39]. These observations suggest variations within *DDX60L* and the interaction between them may contribute to COVID-19 disease severity potentially through altered *DDX60L* involving in COVID-19 hijacking mechanisms.

Super variant ch5_138 (OR=1.306, *p*=5.71×10^−7^) consists of 8 interacting SNPs. In particular, the variant rs41294550 is within the upstream of gene *HSPA9*. The heat shock protein encoded by this gene is important for cell proliferation, stress response, and mitochondrial maintenance. Recent research indicates that gene *HSPA9* knockdown would result in declined B cells in animal models, while memory B cell levels are associated with protection against COVID-19 delta variant infection [40-42]. These data suggest that variations within *HSPA9* may affect the severity upon COVID-19 infection.

Super variant chr6_54 (OR=1.329, *p*=1.67×10^−7^) consists of 4 interacting SNPs, including rs141615401 that lies in the intergenic region of gene *ELOVL5*. The gene *ELOVL5* has been shown to be associated to familial squamous cell lung carcinoma by a previous GWAS study, and several studies indicates that lung diseases would modestly elevate the risk of death after COVID-19 infection [43-45]. In particular, one study notes that patients with lung cancer have more severe symptoms related to COVID-19 infection [52]. Our data suggest that variations within *ELOVL5* and its interaction may result in increased risk of death among COVID-19 patients from lung-related disease.

Furthermore, we capture several SNPs super variants chr7_43 (OR=1.370, *p*=7.61×10^−7^), chr7_151 (OR=1.594, *p*=5.47×10^−9^) and chr10_6 (OR=1.309, *p*=1.12×10^−6^). They are rs7918112 rs78659910 and rs6943608 and their nearby genes are *LASTR, GLI3*, and *AGAP3*, respectively. The genes *LASTR* and *AGAP3* have been reported to be associated with pulmonary function (FEV1/FVC), while the gene *GLI3* has also been found to be related to the pulmonary function (FVC) [46]. COVID-19 virus has been shown to affect many organs, with the lung being one of the most affected [47, 48]. In particular, one study find that severe COVID-19 patients had an abnormal FVC over a half-year following discharge [49]. These findings suggest that future analysis of *LASTR, GLI3*, and *AGAP3* gene expression in lung tissues may provide a better understanding on the molecular pathogenesis of COVID-19.

Super variant chr16_57 (OR=1.394, *p*=1.19×10^−7^) is composed of 9 SNPs. Among them, rs12596894 is inside an intron of gene *NUP93*. Previous studies reported that severe acute respiratory syndrome coronavirus (SARS-CoV) protein nsp1 disrupts localization of *NUP93* from the nuclear pore complex, and the SARS-CoV-2 virus could have the similar disruption on *NUP93* [50]. These observations suggest that a further investigation on the gene *NUP93* may provide a new insight on COVID-19 pathophysiological mechanisms.

Super variant chr20_15 (OR=1.310 *p*=6.40×10^−7^) includes 8 SNPs. They are all located in the intronic regions of nearby gene *MACROD2*. Recent evidence has shown that genetic polymorphisms in the gene *MACROD2* are linked to pulmonary function test parameters [51], and the *MACROD2* is the closest human homolog of COVID-19 with approximately 30% sequence identity and a similar 3D structure [52]. In addition, the *MACROD2* expression is reported to be associated with the removal of mono (ADP-ribosylation), and COVID-19 encodes a nonstructural protein 3 (nsp3) that contains an ADP-ribose phosphatase domain for immune response [53]. Therefore, our results suggest that variations within *MACROD2* is likely associated with COVID-19 related infection and inflammation, potentially due to alterations in the immune response regulating infection and inflammation.

### Comparison with traditional GWAS

We compare super variants identified by DRAG with SNPs identified by traditional GWAS. Again, the same white British ancestry samples with sex, age and 10 PCs are used for GWAS. Through the traditional method, we identify 5 loci (Fig. 2 A) associated with COVID-19 mortality with commonly used threshold of 5 × 10^−8^. We find an association at chromosome 5q32, with the index SNP rs7447800 (*p* = 2.66 × 10^−8^). This index SNP is in the same region as a functional variant in JAKMIP2 gene, specifically rs10477338, which is reported to be associated with testing positive for COVID-19 by a GWAS study conducted on approximately1,000,000 individuals [11]. Fig. 2B shows the location of variants on chromosome 5 identified by both methods and suggests that both strategies should be used in tandem to boost the strength of each method.

**Figure 2:**
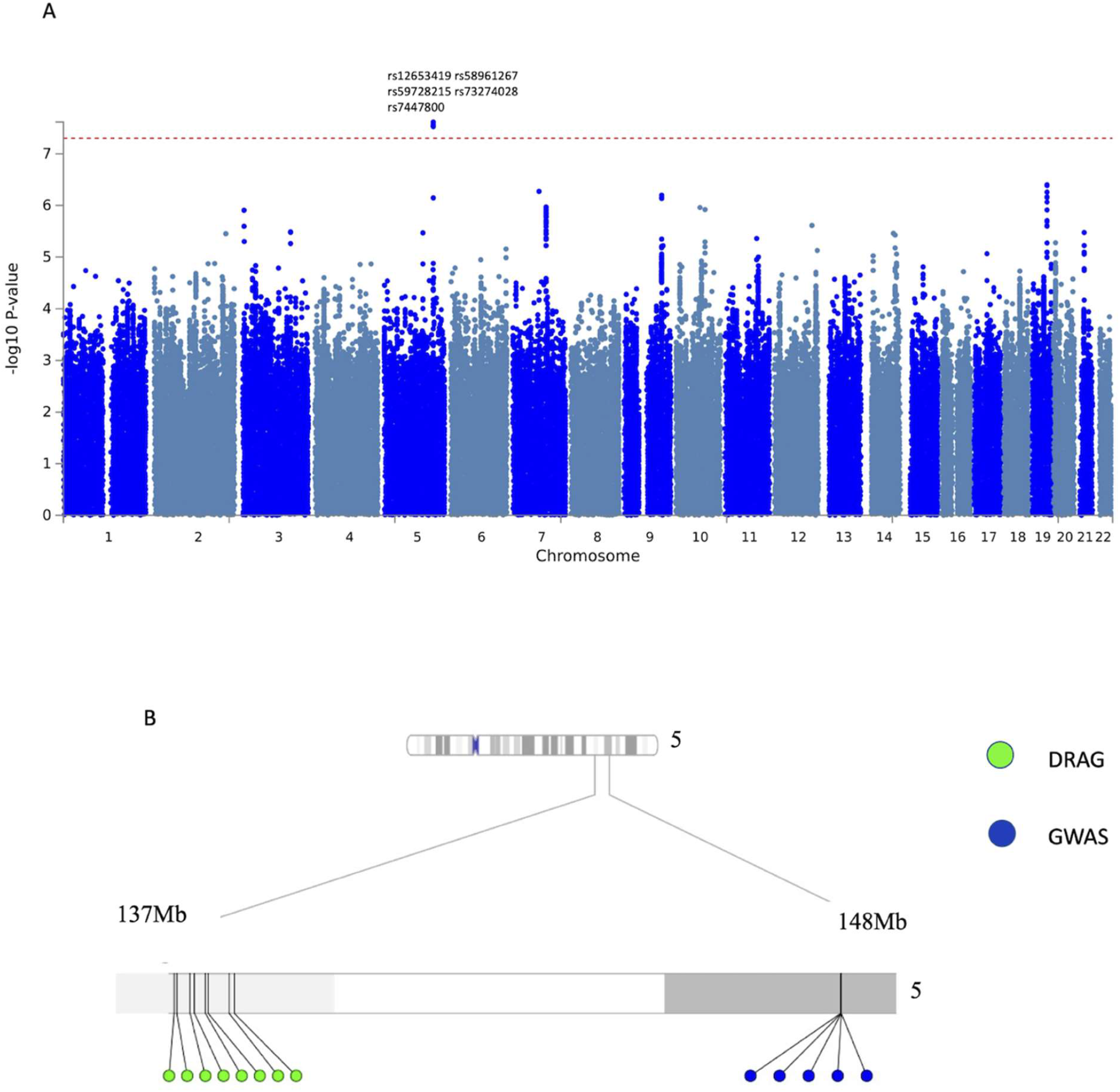
(**A**) Manhattan plot of traditional single SNP association analysis based on samples with white British ancestry only and controlled for gender, age and 10 PCs. The red dashed horizontal line corresponds to the commonly adopted genome-wide significant level at 5 × 10^−8^. (**B**) The location of SNPs on chromosome 5 identified by DRAG (green) and traditional GWAS (blue).

### DRAG increases detection power compared to tree-based model on simulated data

Finally, we perform simulation studies to demonstrate the efficacy of our approach, by comparing it to an established method, tree-based analysis of rare variants (TARV), in various settings. We refer the readers to [20] for more details about TARV implementation. The same procedure for both methods is repeated 200 times in each setting, and the pool of 200 results is summarized in Fig. 3. We evaluate the performance by detection rate, which is defined as the number of correct detections divided by the total number of repeated experiments.

**Figure 3.**
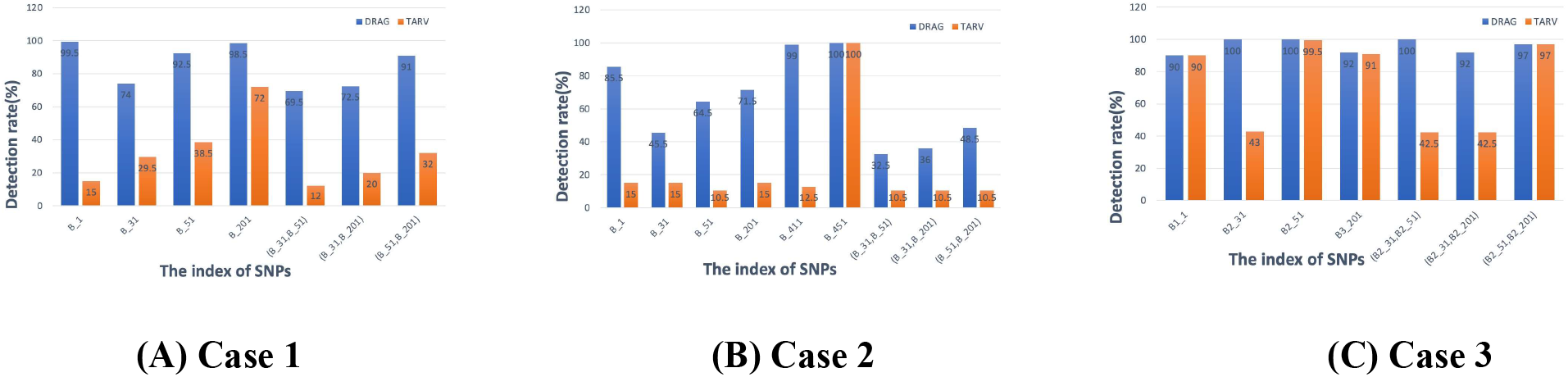
Detection rate of true signal and interaction in different cases. The x-axis of panels (A) through (C) represents the index of SNP. The y-axis of panels (A) through (C) represents the detection rate (%). Blue and red bars represent DRAG and TARV, respectively. The B_k represents the *k*th SNP for first two cases and the Bj_K represents the *k*th SNP in the SNP set *j* for the third case, 1 ≤ *i* ≤ 1,0000, 1 ≤ *j* ≤ 30, 1 ≤ *k* ≤ 500.

Fig 3.A and Fig 3.B exhibit the comparisons between DRAG and TARV in single SNP detection and interacting SNPs detection, respectively. DRAG achieves higher detection rates than TARV in both single SNP detection and interacting SNPs detection. For example, the DRAG on SNP B_411 achieves detection rate of 99% for Case 1, which is better than the TARV on the same SNP (i.e.,12.5%). In addition, DRAG for interacting terms (B_31, B_201) has a detection rate of 32.5% for Case 1 and 69.5% for Case 2, while the TARV only achieves detection rate of 10.5% and 12% respectively, which suggests that DRAG performs better on detecting interactions. Overall, the DRAG outperforms TARV in detecting interacting terms.

Fig 3.C summarizes the performance of DRAG and TARV in the third setting. Both DRAG and TARV work well on detecting individual SNP B1_1 and interactions (B2_51, B3_201), as evidenced by respective similar detection rate for DRAG and TARV of 97% on B1_1 and 97% on (B2_51, B3_201). It is noteworthy that DRAG generates similar results when detecting single SNP B1_1, B2_31 and B2_51 as TARV. However, DRAG on single SNP B2_31 achieves detection rate of 100%, which is significantly superior to the TARV (i.e., 43%). Similarly, as with the DRAG, the observed detection rate for interaction term of (B2_31, B2_51) and (B2_31, B3_201) (100% and 92%, respectively), are superior to TARV from (B2_31, B2_51) with 42.5% and (B2_31, B3_201) with 42.5%. The results suggest DRAG outperforms the TARV by a large margin, which implies that deep learning-based model is superior to tree-based model at detecting complex interacting SNPs when large amounts of data are presented.

## Discussion

In summary, we train deep learning to learn the association between genetics variants and COVID-19 related mortality using genotyping data from the UK-biobank white British cohort. We identify 15 super variants and explored their relationships to COVID-19 related mortality. We also report 54 genes related to detected super variants, many of these genetic variants overlap with previously reported associations with lung-related phenotypes or COVID-19 outcomes or inflammatory diseases.

In addition to novel genetic discoveries in COVID-19, our approach is also novel in several statistical aspects and possesses unique strengths: the ranking and aggregation strategy by an iterative super variant search enables us to capture complex non-linear interacting SNPs and consequently and obtain group of interacting SNPs with high COVID-19 related mortality risk potential. In contrast to the classic CAE approach, the DRAG incorporates two innovative processes: iteratively running CAE procedure and model selection through BIC. These steps enable the user to explore various combinations of SNPs and select the optimal one. Finally, we demonstrate in simulation study that DRAG is superior to tree-based model by a significant margin when we detect complex interacting SNPs that are entangled in high-dimensional nonlinear representations. We believe this is a significant advantage of our strategy since host genetic factors that account for disease risk frequently include multiple complex interactions. Together, our findings provide timely clues and prospective directions for better understanding the molecular pathogenesis of COVID-19, which may have implications for clinical treatment of this disease.

## Limitations

Our study is subject to several limitations. The study population largely consisted of white British ancestry UK Biobank participants, limiting generalizability to other populations. Because only about 5% of the UK Biobank population had COVID-19 data available, it will be interesting to explore our findings when additional data and populations of diverse ancestry become available. Although one can vary the degrees of non-linearity and flexibility in the model by tunning hyper-parameters like *K*_0_, learning rate and the number of layers, etc., the computation cost of DRAG would increase dramatically as the value of *K*_*0*_ increases. We expect to implement parallel computation in future work to make our procedure more user-friendly. Finally, the roles of the identified super variants and associated genes are not substantiated by functional validation. Still, our findings warrant future investigation to learn the associations between genetic variants and the COVID-19 outcomes.

## Method

### Data and preprocessing

#### Cohort dataset

The data used in the preparation of this study were obtained from the UK Biobank (project ID: 42009) [54, 55]. The UKB was launched in 2006, led by Heart Foundation Professor Sir R. Collins. The primary goal of UKB has been to help researchers to better understand a range of common and life-threatening diseases. The UKB cohort recruited 500,000 individuals aged 40–69 in the UK via mailer from 2006 to 2010. In total, we analyze 148,654 participants with COVID-19 data (Field ID: 40100) as of late December 2022. Out of these participants, 28,652 (19.27%) had the positive polymerase chain reaction (PCR) test result.

#### Data preprocessing

We first discard the UKB samples for which age or sex was missing. Second, we remove samples identified as outliers in heterozygosity and missing rates. We then delete samples which were identified as putatively carrying sex chromosome configurations that are not either XX or XY. These were identified by looking at average log2Ratios for Y and X chromosomes. Next, we remove the participant who are excluded from kinship inference process. Finally, we only consider white British participants to limit the potential effect of population structure. After standard sample quality controls, there remain 28,097 of COVID-19 infected participants in our final analyses, of which 1,656 are deaths (5.89%) and 26441 are survivors (Fig 4.A).

**Figure 4:**
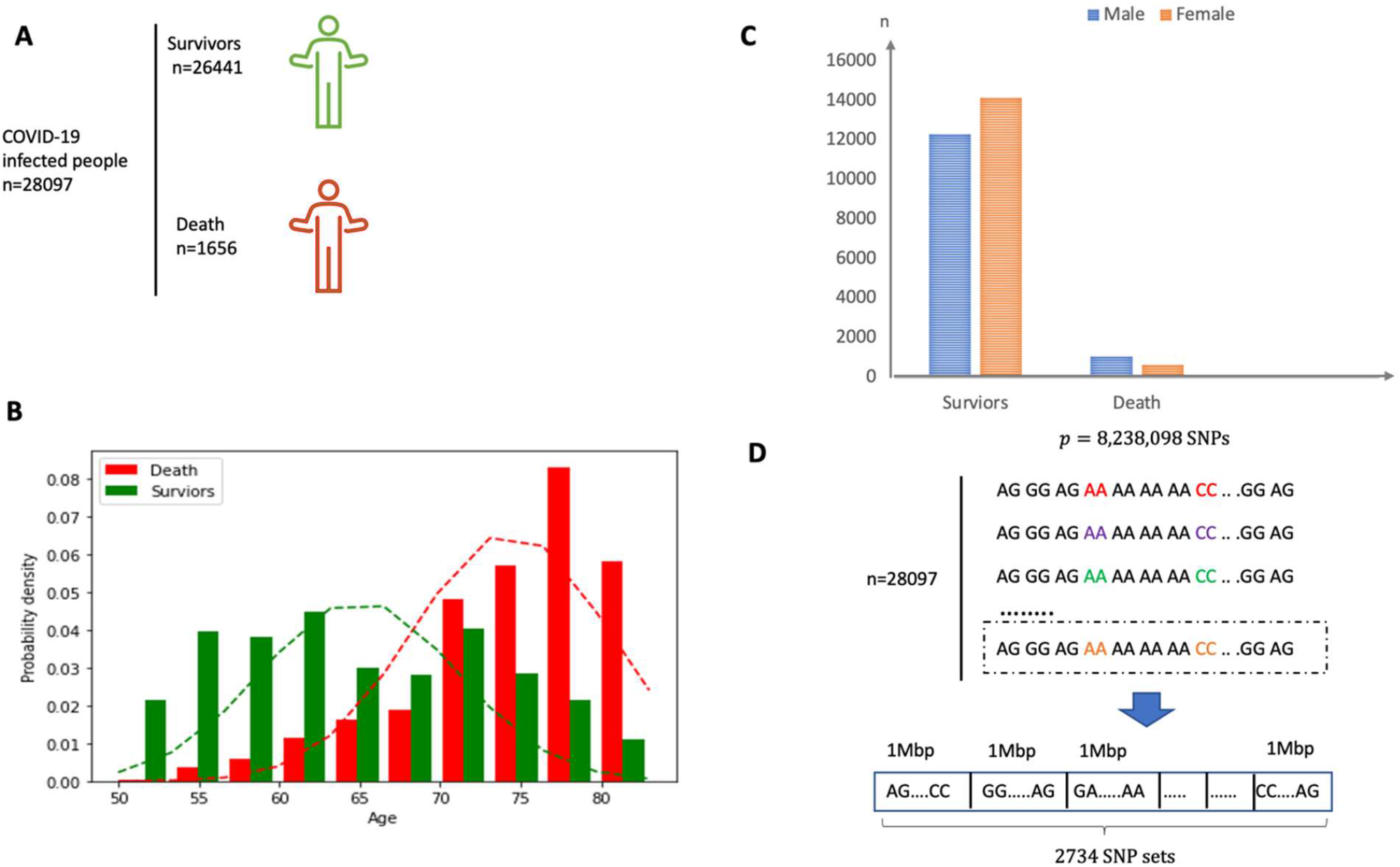
**(A)** Overview of the participants included and the samples and data collected. **(B)** Sex distribution in both survivor and death group. **(C)** Age distribution in both survivor and death group. The mean of age for death group is around 75 years old. **(D)** The SNP dataset are divided into 2734 non-overlapping local sets according to the physical position and each set consists of SNPs within a segment of physical length 1 Mbp.

#### Definition of Phenotype

To detect the genetic variants associated with the risk of COVID-19 related mortality, we define a death as a person who died due to COVID-19 infection and a survivor as a person who had the positive polymerase chain reaction (PCR) test result but survived.

#### Demographic variables

Age and sex (Field ID: 31, 34) are used as confounding variables in regression analyses to eliminate potential bias when testing the significance of association between super variants and the phenotype (Fig 4 B&C). For sex, we used the genetic sex when available, and the self-reported sex when genetic sex was not available.

#### Genotype data and quality control

We use imputed genotyping data (Field ID: 22801-22822) from UK Biobank as genetic predictors in the present study. SNPs with duplicated names and positions are deleted. In addition, we remove variants with minor allele frequency (MAF) ≤ 0.05 and disrupted Hardy-Weinberg equilibrium (p value *<* 10^−6^). We retain in total 8,238,098 SNPs. We next divide the whole SNP dataset into 2734 non-overlapping local SNP-sets according to the physical position so that each SNP-set consists of SNPs within a segment of physical length 1Mbp (Fig 4.D).

### Study design

All analyses are conducted in the UK Biobank unless otherwise stated. We first adopt the concept of super variants. In contrast to a gene, which refers to a physically connected region of a chromosome, the loci contributing to a super variant can locate in any part of the genome. Since the super variant aggregates the strength of distinct signals, it has been shown that it is both powerful and stable in association studies [17, 18]. Furthermore, super variants take into consideration the potential complex interactions among loci.

Our design considers the following discovery-validation procedure (Fig. 5). The whole dataset is partitioned into two sets, one for discovery and the other for verification with ratio of 2:1. A total of 1,104 fatalities and 17,627 survivors are included in the discovery set, whereas 8,814 deaths and 552 survivors are included in the verification set. We train the DRGA to find the initial candidate super variants in the first half of discovery set, the logistic regression is then applied to the second half of the data from discovery set to find initial optimal super variants, permitting us control for Type I error. The SNPs contributing to the initial optimal super variants are extracted and aggregated into super variants on the verification datasets. We validate their associations with the COVID-19 related mortality through logistic regression on the verification set. We use 1.83× 10^−5^ (i.e., 0.05/2734, adjusted for the number of super variants) as the Bonferroni-corrected cut-off p-value for one super variant on the discovery dataset. A super variant is verified if its logistic regression coefficient achieves the level of 0.05 significance on the verification dataset. Statistical analyses were conducted with Python 3.9.0.

**Figure 5.**
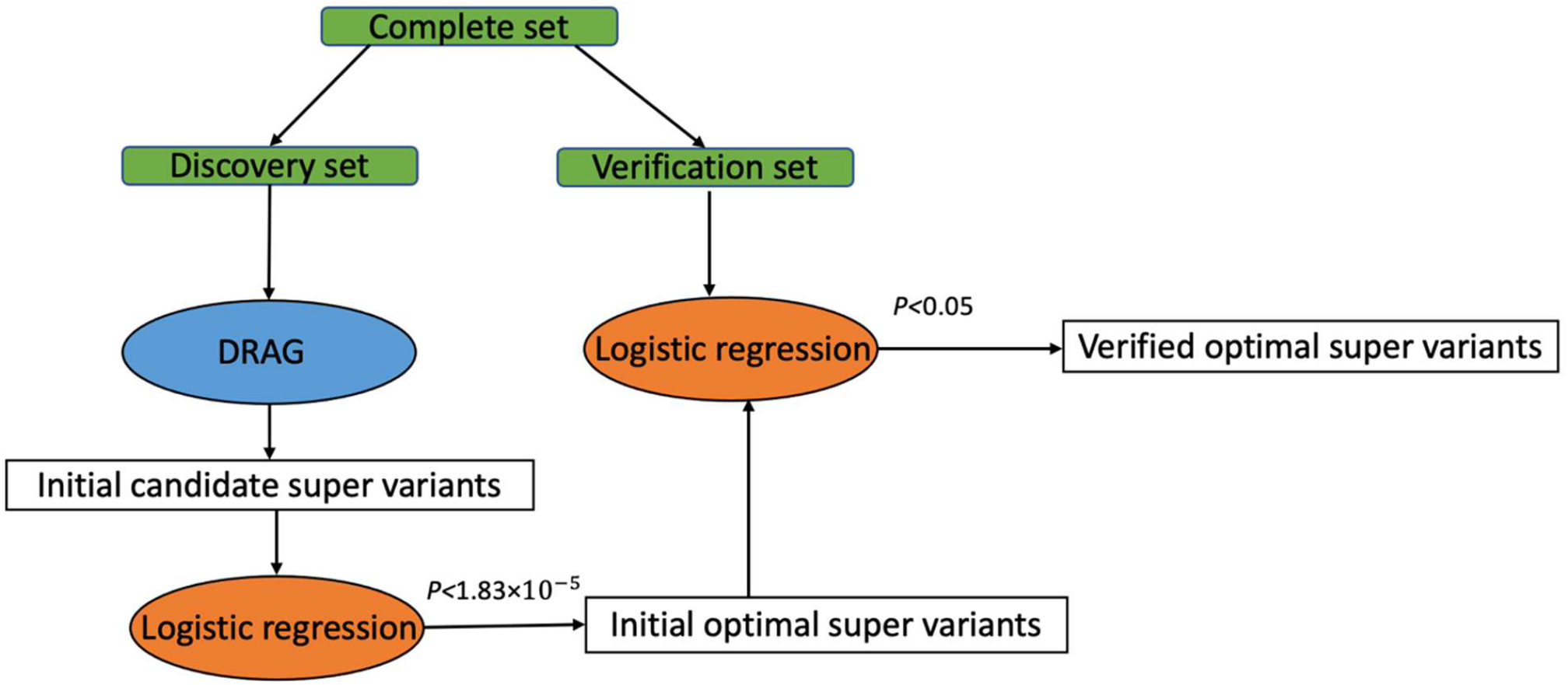
Overview of discovery-validation procedure. Complete dataset is partitioned into two sets, one for discovery and the other for verification. Discovery set is used to find optimal super variants and verification set is used for evaluating the selected super variants.

### Machine learning algorithms

#### Overview of DRAG algorithm

DRAG algorithm consists of three main steps as summarized in Fig 6. In the first step, chromosomes are divided into different non-overlapping local genomic sets according to their physical positions. In the second step, the CAE is utilized within each set to select a subset of k SNPs, where k is user-specified and spans a certain range, e.g., (1, *K*_*0*_), *K*_*0*_ is a hyper-parameter. For each chosen k, we train a CAE model. Later, these CAE models will be evaluated based on some criteria, and a best k will be picked. In another words, *K*_*0*_ different subsets of SNPs are selected for each genomic set by iteratively training CAE. In the third step, a super variant is formed by a subset of SNPs that indicates whether any minor allele is present [16]. A total of *K*_*0*_ super variants are formed. We use logistic regression combined with BIC to identify the optimal super variant among the *K*_*0*_ super variants with each genomic set. The optimal super variant is the set of SNPs that minimizes the BIC value.

**Figure 6:**
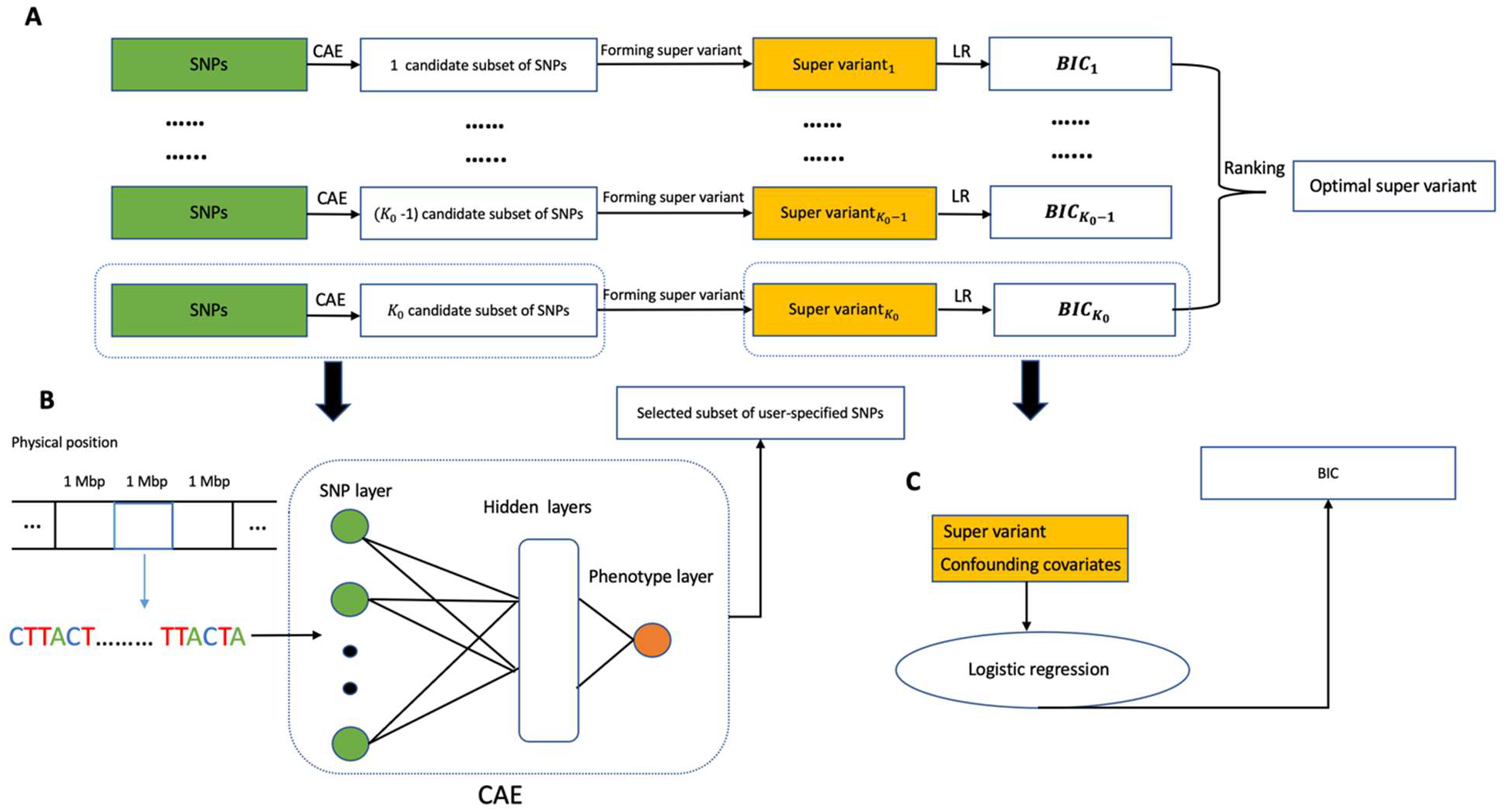
Visual representation of the proposed SNP selection approach using DRAG. **(A)** An overall flowchart of the approach for selecting of super variant. **(B)** CAE model training and finding the selected subset of user-specified SNPs. **(C)** Logistic regression training and finding BIC value.

#### Concrete Auto-encoder architectures

We design a deep neural network architecture to learn complex relations between genetic variants and phenotypes (Fig 7.A). Let vector ***x*** ∈ *R*^*p*^ be a collection of SNPs and *y* be a phenotype which is a binary random variable taking value 0 or 1. We first use a user-specified set of *k* SNPs denoted by ***z***. We have the ***z*** = ***B***^*T*^***x***, where ***z*** ∈ *R*^*k*^and ***B*** ≡ [***β***_1_,.., ***β*** _*k*_] ∈ *R*^*p×k*^ denotes the parameters between SNPs layer and user-specified subset layer. Thus, *z*_*i*_ = ***β***_*i*_^*T*^*x* = *x*_*1*_ *β*_*i1*_+…+*x*_*p*_*β*_*ip*_, *i* = 1, …, *k*, is referred to as to the *ith* nodes in user-specified subset layer. Decoder layers consist of hidden layers and phenotype layers and serve as the reconstruction function. Decoder layers take the outputs ***z*** from user-specified subset as its inputs. A decoder with *h* hidden layers is defined as *f*(***z***) = *f*^*(h)*^(…*f*^2^(*f*^1^(***z***))), where *f*^*(i)*^(·) represents the *ith* hidden layers and the output *f*(***z***) consists of the predicted phenotype for an individual. The loss function for binary classification is given by:

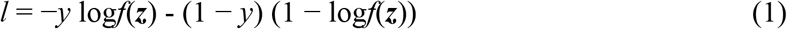

**Figure 7:**
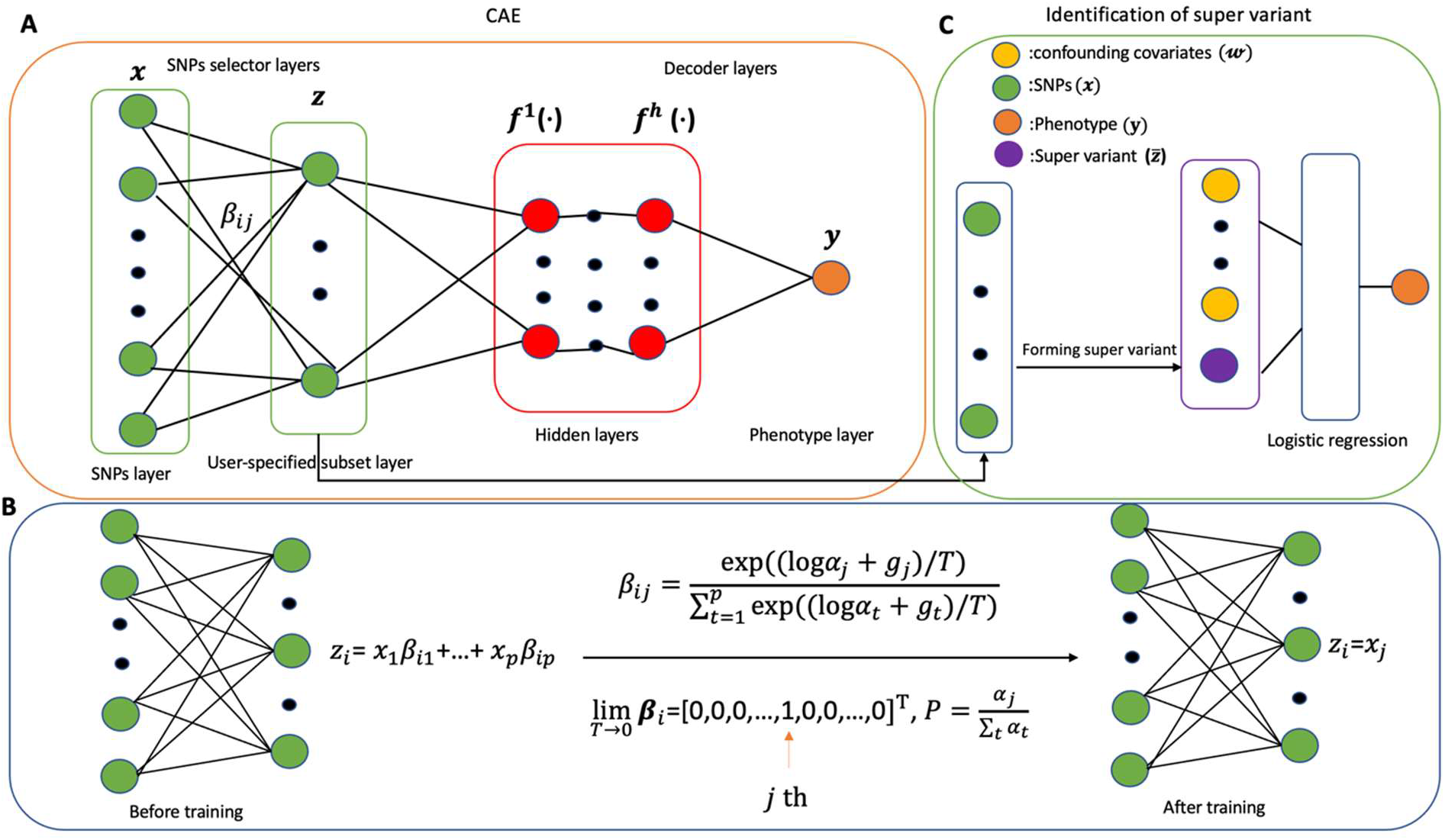
**(A)** An overall of representation of the Concrete auto-encoder approach. *x* and *y* are the set of SNPs and the phenotype, respectively. *z* denotes the set of top identified SNPs and *f*^(*i*)^(*·*) represents the outputs in each hidden layers. **(B)** A mathematical overview of SNP selection procedure using CAE on SNPs selector layers. **(C)** The identified SNPs from (A) and confounding covariates are then used for logistic regression to facilitate the identification of super variants.

In our specific application, we let *h* = 2, *n*_1_ and *n*_2_ denote the number of nodes in each layer, and 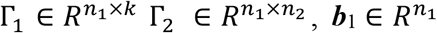 and 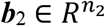 denotes the weights and bias term attached to each hidden layer *j* (*j* = 1,2). Then the outputs in each hidden layer are *f*^1^ (***z***) = *ϕ*_1_(Γ_1_ ***z*** +***b***_1_) and *f*^2^ (***z***) = *ϕ*_2_(Γ_2_ *f*^1^ (***z***)+ ***b***_1_), where *ϕ*_*j*_ are activation functions of each layer. In particular, the activation functions for the hidden layers are ReLU: *ϕ*(*x*) = max (0,*x*). The predicted 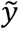 can be written as 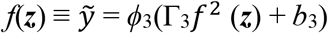, where *ϕ*_3_ is the activation function, which is typically a logistic function for dichotomous traits, Γ_3_ ∈ R^*n*2^ and *b*_3_ are the weights and bias in phenotype layer. Therefore, the optimization function for the data under the truth can be written as:

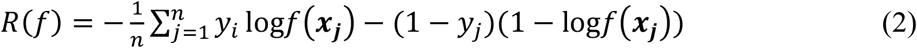

where *n* is the total number of subjects and *x*_*i*_ and *y*_*i*_ are set of SNPs and phenotype of *ith* subject, respectively.

In practice, the user-specified subset layer generally outputs weighted linear combination of input variants. We next describe how the user-specified subset layers works, that is, how each variant is selected (Fig 7.B). To preform the variant selection, the element of ***β*** has be to either 0 or 1. We now only consider *ith* node in user-specified subset layer and sample *p*-dimensions vector ***β***_*i*_ using the proposed technique in [23, 56, 57]:

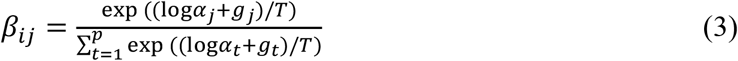

where *β*_*ij*_ refers to the *jth* element in ***β***_*i*_, *T* ∈ (0,∞) is the so-called temperature parameter, *α* ∈ *R*^*p*^ is a p-dimensional vector which all the elements are strictly greater than zero and *g* ∈ *R*^*p*^ is a p-dimensional vector which all elements are generated from a Gumbel distribution [58]. In this way, we obtain the ***β***_*i*_ smoothly approaches the discrete distribution as *T* ⟶ 0, outputting one hot vectors with *β*_*ij*_ = 1 with probability 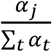. That is 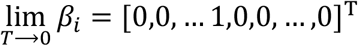. Therefore, the *ith* node in user-specified subset layer outputs exactly one of the variants when *T* ⟶ 0. Using the same logic we sample a p-dimensional random vector ***β*** for each of the *k* nodes to perform variant selection. Note, for the initialization of *T*, Abid [23] claimed that the model is unable to explore different feature combinations and converges to a poor local minimum if the initial value of *T* is set to be extremely low. We apply their work and use the varying temperature *T*(*e*) = *T*_1_(*T*_2_*/T*_1_)^*e/E*^, where *T*(*e*) is the temperature at epoch number e, and *E* is the total number of epochs to train the model, *T*_1_ and *T*_2_ are tuning parameters which are usually set to be a high value for *T*_1_ and a low value for *T*_2_. With such a choice of ***β***, the optimization function (2) can be minimized with respected to ***α***, **Γ**_*j*_ and ***b***_*j*_ (*j* = 1,2,3) using the backpropagation algorithm and the user-specified set of *k* SNPs are selected afterwards.

#### Identification of super variants through logistic regression

Since the optimal number of SNPs remains unknow, the super variants learned by CAE may be sub-optimal. To resolve this issue, we use a model selection technique with BIC in logistic regression, in which the target is the phenotype and the main effect estimated by super variant that consists of selected SNPs (Fig 7.C). To facilitate the identification of super variants, all possible values for the number of nodes in user-specified set layer with observations ***z*** are inspected using logistic regression. The range of values to consider for the *k* is greater than 1 and less than *K*_*0*_, where *K*_*0*_ ≤ *p* and *p* is the total number of variants. For each candidate value *k* ∈ [1, *K*_*0*_], the model with smallest BIC value is selected and the corresponding variants aggregated to form the optimal super variants. As an illustration, let *z*_*1*_,…*z*_*k*_ be the variants selected by CAE and 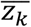 denote super variant and *w*_*iq*_ denote the *qth* covariate of subject *i*. A general model is given by

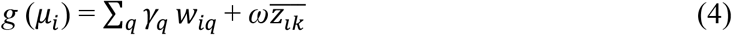

where *μ*_*i*_ = *E* (*y*_*i*_), *y*_*i*_ is the phenotype for subject *i*, and *g* is a link function which is logit function for binary classification, 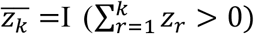, and *γ*_*q*_ and *ω* are coefficients for covariates and genetic variants, respectively. We next apply model (4) on formed super variant and confounding variables to find the BIC value denoted by *BIC*. The importance of each model can be evaluated based on the magnitude of *BIC*. Finally, we choose the super variant which consists of *k*_*b*_ variants 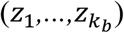 with the smallest *BIC* as the optimal super variant, where 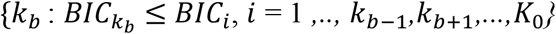

#### Tuning

There is a negligible impact of tunning parameters, *K*_*0*_, learning rate, the number of layers and the number of nodes in each layer on model performance. Especially, the value of *K*_*0*_ is critical for the success of our algorithm. The range of values to consider for the *K*_*0*_ for each set is no more than the total number of SNPs in this set. Too large a *K*_*0*_ would result in unnecessary computation burden, while too small a *K*_*0*_ might produce a sub-optimal super variant. In this study, we choose *K*_*0*_ to be 25 and use 128 and 64 nodes to each hidden layer of 2-hidden layers CAE with the Adam optimizer of learning rate of 0.001, which is tuned on an independent identically distributed dataset.

### Simulation framework

For the simulated data set up, we use the genotype data on Chromosome 22 from the UK Biobank COVID-19 dataset because there is no significant signal identified on this chromosome in previous studies. We choose 10,000 patients at random. For the first two cases, we randomly sample 500 SNPs from a single SNP set to form the synthetic genetic dataset. In the third case, we randomly select 30 SNP sets and 500 random SNPs from each set to create the whole synthetic genetic dataset. As a consequence, a super variant discovered by both DRAG and TARV is considered to be significant if the p-value is less than 0.05 (Case 1 and 2) and 0.05/30 (Case 3), respectively. The disease *y* is generated from *y*_*i*_ *∼* Bernoulli (*p*_*i*_), and

**Case 1:** 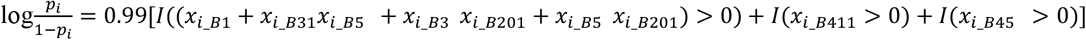

**Case 2:** 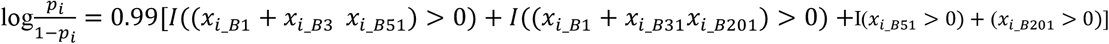

**Case 3:** 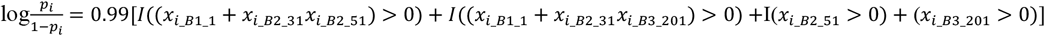

where *x*_*i_Bk*_is the *k*th SNP for subject *i* for first two cases and *x*_*i_Bjk*_ represents the *k*th SNP in SNP set *j* for subject *i* for the last case, 1 ≤ *i* ≤ 1,0000, 1 ≤ *j* ≤ 30, 1 ≤ *k* ≤ 500. Case 1 includes individual effect expressed as B_411 and B_451. In addition, both cases consist of same interaction term of (B_31, B_201) a (B_31, B_51) and (B_51, B_201). There are a total of 6 true signals in Case 1 and 4 true signals in Case 2. We consider an interaction term being identified if all true signal SNPs within the same set, such as the two SNPs B_31 and B_51 are identified for case 1 at the same time. For brevity, Case 3 consists of 4 true signals from three different SNP sets with two different structures, individual signal, interacting signal with group sizes 2.

## Data Availability

Data are available from the UK Biobank.

https://www.ukbiobank.ac.uk/

## Data availability

UK Biobank data is made available to researchers from universities and other research institutions with genuine research inquiries, following the UK Biobank approval. (https://www.ukbiobank.ac.uk).

## Author contributions

HZ conceived and oversaw this project, ZL and WD took the main responsibility in execution of this project, SW and YY contributed to the data analysis. All authors made critical input to the manuscript.

## Acknowledgments

Zhang’s research is supported in part by U.S. National Institutes of Health (R01HG010171 and R01MH116527) and National Science Foundation (DMS-2112711). This research has been conducted using the UK Biobank Resource under Application Number 42009. We thank the Yale Center for Research Computing for guidance and use of the research computing infrastructure.

## Supplementary information

**Table S1:**
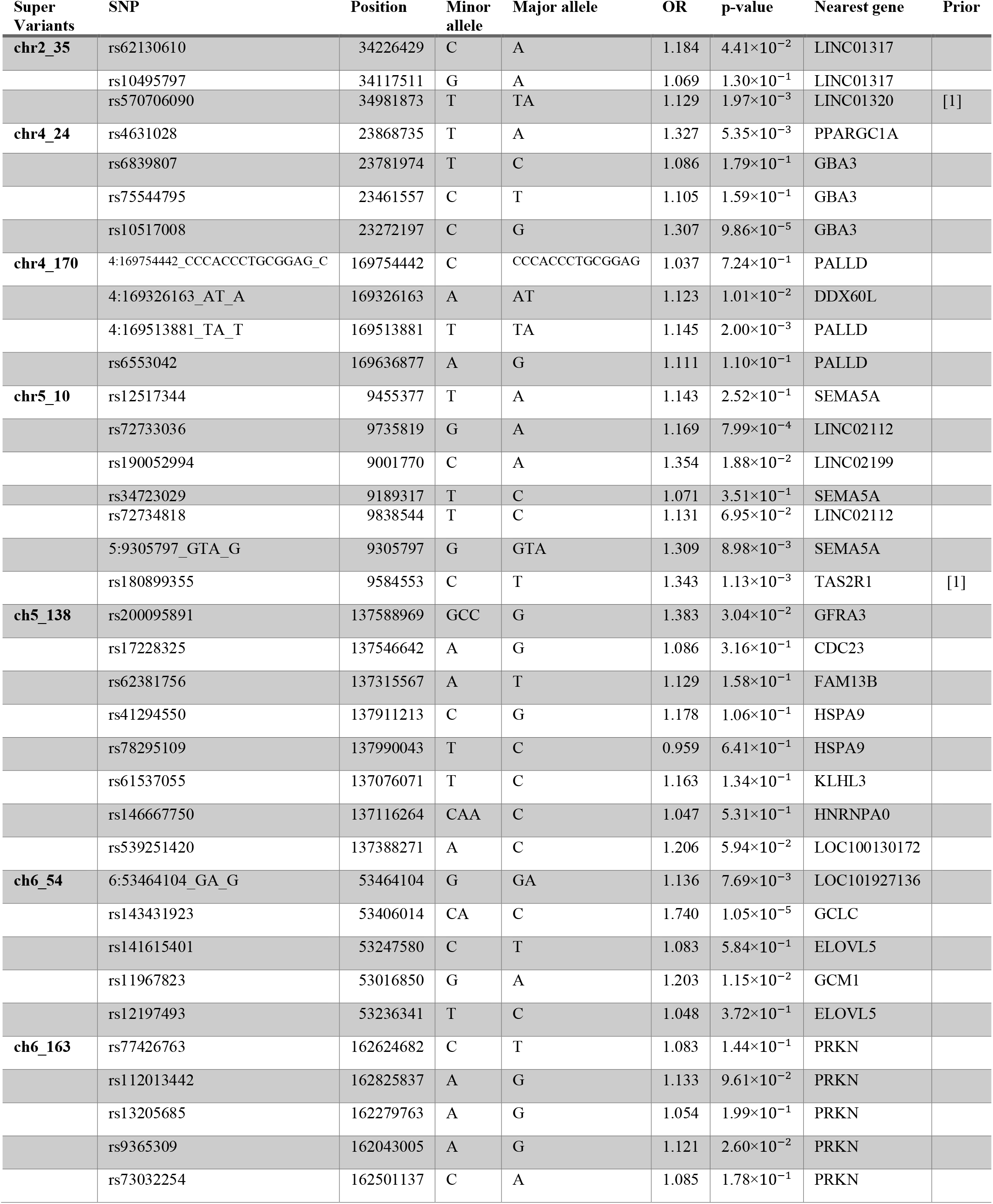
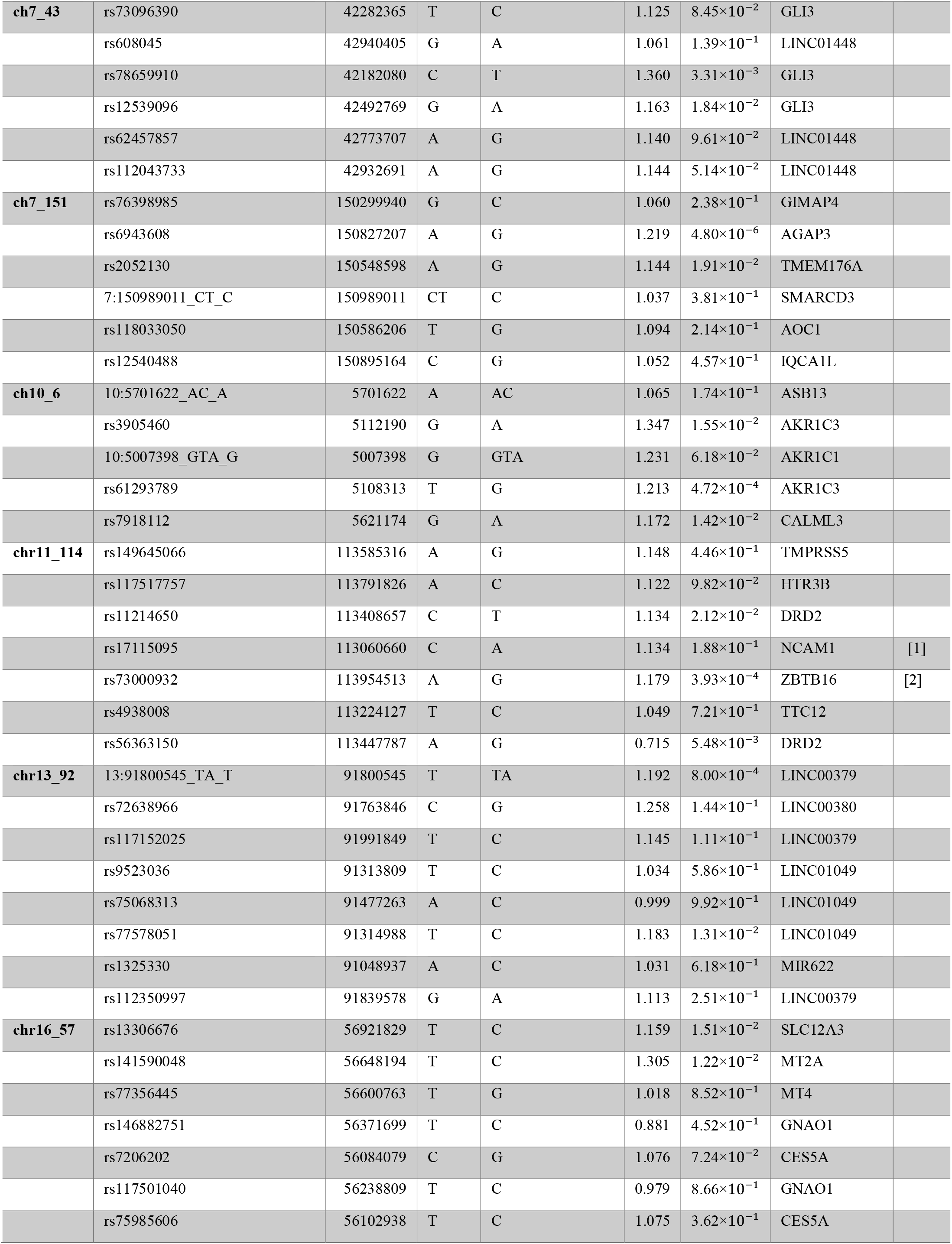
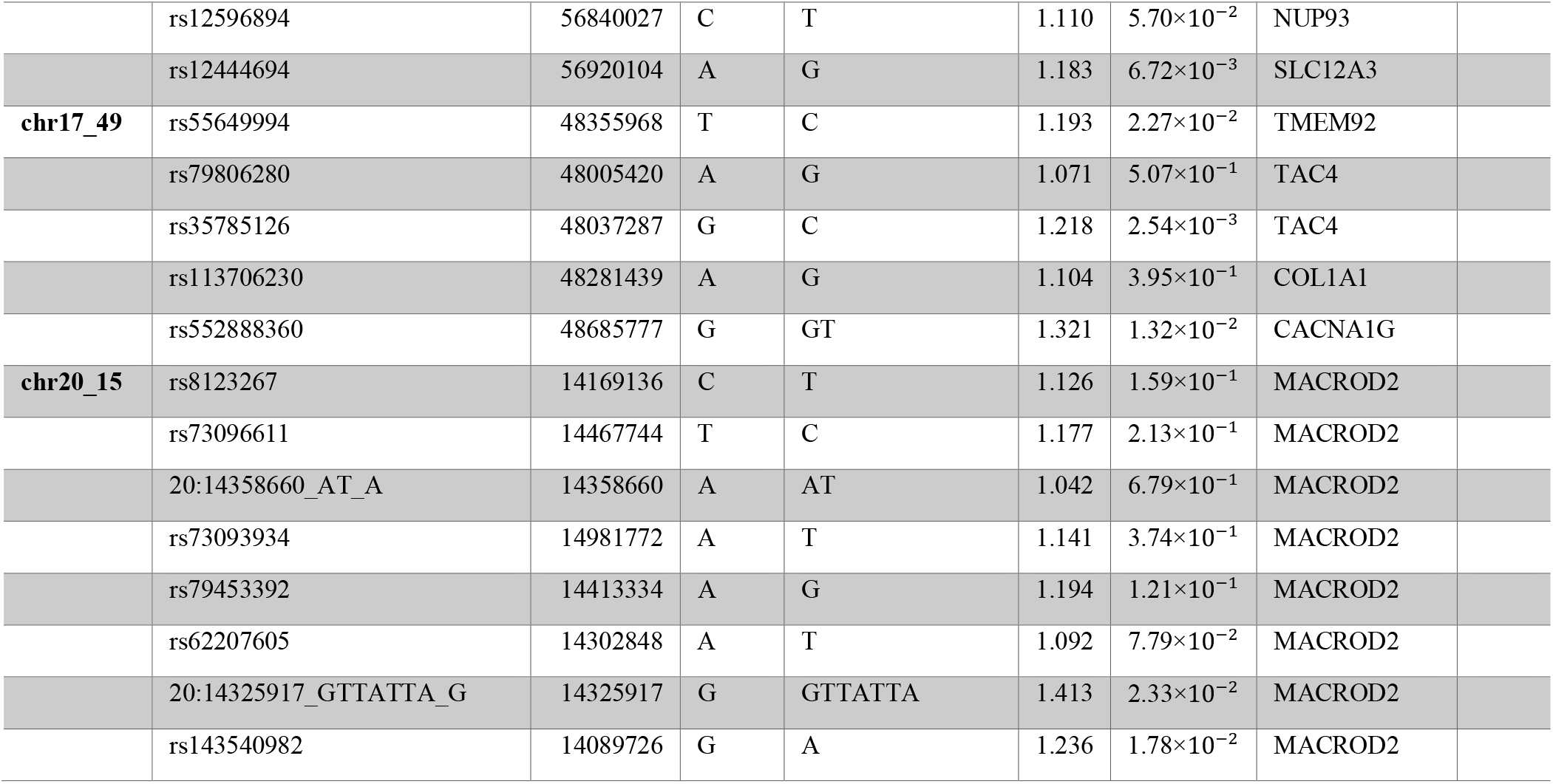
Corresponding SNPs in verified super variants with DRAG. The chr i_j represents the super variant in the *j*th set of the *i*th chromosome; SNP, the rsID of the variant, where available; Prior, known from prior publications addressing common genetic variation linked to COVID-19; OR: odds ratio.

